# Combination of Baricitinib plus Remdesivir and Dexamethasone improves time to recovery and mortality among hospitalized patients with severe COVID-19 infection

**DOI:** 10.1101/2022.04.04.22273425

**Authors:** Victor Perez-Gutierrez, Virali Shah, Afsheen Afzal, Amnah Khalid, Ariane Yangco, Sebastian Ocrospoma, Nail Cemalovic, Anjana Pillai, Moiz kasubhai, Vihren Dimitrov, Vidya Menon

## Abstract

**Background:** There seems to be a gap in the therapeutic options for severe Covid-19 pneumonia. Though the beneficial effect of combination treatment with baricitnib and remdesivir in accelerating clinical status improvement is described, the impact of the triple therapy with baricitinib + remdesivir/dexamethasone is not known.

**Methods:** A retrospective observational study comparing the effect of baricitinib plus standard treatment (remdesivir and dexamethasone) with standard therapy in patients requiring ≥ 5 L/min O2 was conducted. The primary outcome was to compared time to recovery in both groups, and the secondary outcomes was to determine mortality rate at discharge.

**Results:** Of 457 patients hospitalized during the study period, 51 patients received standard treatment while 88 patients received baricitinib plus standard treatment. In baricitinib group, the rate ratio of recovery was 1.28 (95%CI 0.84-1.94, p=0.24) with a reduction in median time to recovery of 3 days compared to standard treatment group. Subgroup analysis based on Ordinal Scale showed reduction in median time to recovery by 4 and 2 days with rate ratio of recovery of 2.95 (1.03-8.42, p =0.04) and 1.80 (1.09-2.98, p=0.02) in Ordinal Scale 5 and 6 respectively. No benefit was found in the Ordinal Scale 7 subgroup. An overall decrease in rate (15.9% vs 31.4% p=0.03) a likelihood (OR 0.41, 95%CI 0.18-0.94, p=0.03) of mortality was observed in the baricitinib group. Bacteremia and thrombosis were noted in the Baricitinib group, but comparable with the Standard of care group.

**Conclusion:** Baricitinib with standard therapy reduced time to recovery and offer mortality benefit in patients with severe COVID-19 pneumonia.”

## Introduction

Infection due to SARS-CoV2 characteristically ranges from no symptoms to critical illness, with close to 30% of patients meeting criteria for Acute Respiratory Distress Syndrome (ARDS)[1,2]. In-hospital mortality has been declining during the course of the pandemic, with survival improving from 74.4% (March 2020) to 92.4% (August 2020)[3,4]. However, with the advent of the variants, the risk of mortality has increased, as shown by a 64% higher risk of 28-day mortality due to the B.1.1.7 variant in people older than 30 years [5]. The options of treatment are evolving rapidly, with new/repurposed drugs approved by US FDA and drugs made available under FDA Emergency Use Authorization for treatment of COVID-19 infection[6].

The present understanding of SARS-CoV2 infection is that, on viral binding, ACE2 release via cleavage by ADAM17 (metallopeptidase domain 17) results in activation of IL-6 receptor-mediated activation of STAT3 (Signal transducer and activator of transcription 3), thereby inducing a full activation pf NF-kB (nuclear factor kappa B) pathway, leading to multiple inflammatory pathologies[7]. The role of IL-6 in SARS-CoV-2 induced inflammation suggests that Janus Kinase (JAK) inhibitors may be an attractive therapy for severe COVID-19 patients[7,8].

The options of treatment for severe and critical COVID-19 infection are limited at present. The RECOVERY study demonstrated that dexamethasone resulted in lower 28-day mortality among those receiving oxygen alone or invasive mechanical ventilation[9]. National Institute of Health recommendations include using remdesivir with dexamethasone for hospitalized patients with COVID-19 who require supplemental oxygen[10]. The use of intravenous methylprednisolone also demonstrated to have mortality benefit in patient hospitalized for severe Covid-19 infection[11]. The REMAP-CAP and RECOVERY studies have shown that tocilizumab improved survival, especially in critically ill COVID-19 patients receiving organ support[10,12]. Based on the ACTT-2 (Adaptive COVID-19 Treatment Trial-2), which showed that baricitinib (orally administered JAK1/2 inhibitor) with remdesivir was superior to remdesivir alone in improving oxygenation and clinical status among patients receiving high flow oxygen and non-invasive ventilation, the FDA granted a EUA for the use of baricitinib in combination with remdesivir in November 2020[13,14]. Combination of baricitnib with corticosteroids have also shown an improvement in oxygen requirement and lung functions[15,16]. Here we report the preliminary results of a retrospective observational study of the combination of baricitinib plus remdesivir and dexamethasone with remdesivir and dexamethasone in adult patients hospitalized with severe and critical COVID-19 infection.

## Methods

### Study design and patient population

This a case-control retrospective study in a New York City Health + Hospital acute care facility in the South Bronx, approved by the Institutional Review Board [IRB # 21-007]. We included all adult patients hospitalized with a positive SARS-CoV-2 RT-PCR, radiology features of COVID-19 pneumonia, and requiring ≥5 L/min of supplemental O2. The hospital protocol recommends using dexamethasone 6 mg daily with remdesivir (Veklury®) 200 mg loading, followed by 100 mg daily as standard therapy for the duration of oxygen requirement not exceeding 10 days. After January 1, 2021, the hospital protocol recommended the addition of baricitinib (Olumiant®) administered at a dose of 4mg daily with remdesivir for a maximum of 10 days for all patients requiring ≥5 liters of oxygen. Patients older than 75 years or with creatinine clearance between 30-60 ml/kg/min received 2mgs of baricitinib with remdesivir. Exclusion criteria included baseline chronic oxygen supplementation, CHILD C cirrhosis, AIDS, CKD greater than Stage 3, active cancer, tuberculosis, viral hepatitis, use of DMARDS as methotrexate, cyclophosphamide, azathioprine, mycophenolate mofetil, chronic use of corticosteroids (>10 mg/day), DVT in the last three months and pregnancy. Negative procalcitonin, blood, urine cultures, with ferritin-to-procalcitonin ratio ≥877 (Gharamti et al.) was used to screen for superimposed bacterial in patients on Baricitinib with standard of care regimen. In case of suspicion of superimposed bacterial infection, baricitinib was not used [17].

Two different periods were identified for the inclusion of patients. Patients who fit the inclusion criteria from July 1 to December 31, 2020, served as the “control” group and had received dexamethasone and remdesivir as standard care. Meanwhile, qualifying patients from January 1 to February 20, 2021, who received baricitinib in addition to remdesivir and dexamethasone as part of treatment protocol represented the “case” cohort. All patients were followed up till discharge/death or until March 4, 2021, if they were still hospitalized.

### Data collection and definition of variables

Sociodemographic and broad characteristics including age, sex, race, body mass index, presence of comorbidities (HTN, diabetes, COPD, asthma, coronary artery disease, congestive heart failure, previous history of cancer, HIV infection, rheumatologic disease, and former smoker) were obtained by electronic chart review. Charlson Comorbidity Index was calculated to estimate the intrinsic aggregate comorbidity burden[16]. Clinical data, including vital signs on admission, were obtained. Pulse oximetric saturation (SaO2/FiO2) (S/F) ratio was substituted for the PaO2/FiO2 ratio to assess acute lung injury’s oxygenation criterion[19]. Sequential Organ Failure Assessment (SOFA) was measured within 24 hours of the requirement of mechanical ventilation, or the highest oxygen demand during the hospital course[20]. An eight-category Ordinal Scale was used to assess daily clinical status from admission today of recovery [13]. The severity of Covid-19 infection on admission was stratified based on the eight-category Ordinal Scale. Moderate illness corresponded to ordinal scale 4, which represent no requirement of oxygen supplementation. Severe illness if the Ordinal Scale was 5 or 6, which represent patients requiring low flow and high flow oxygen system, respectively. Critical illness if the Ordinal Scale was 7, meaning that endotracheal intubation was performed during the day of admission. This stratification can be considered as surrogate of the National Institute Health guideline with last issued on April 21, 2021, witch define Moderate illness as individuals who show evidence of lower respiratory disease during clinical assessment or imaging and who have an oxygen saturation (SpO_2_) ≥94% on room air at sea level; severe illness as Individuals who have SpO_2_ <94% on room air at sea level, a ratio of arterial partial pressure of oxygen to fraction of inspired oxygen (PaO_2_/FiO_2_) <300 mm Hg, respiratory frequency >30 breaths/min, or lung infiltrates >50%; and critical illness as individuals who have respiratory failure, septic shock, and/or multiple organ dysfunction[21].

### Outcome

Day of Recovery was defined as the day of discharge, or the day when the patient required less than 3 liters of supplemental O2 with a National Early Warning System Score of <=2 for 24 hours or no longer needed oxygen supplementation or medical care[13,22].

### Statistical analysis

Intergroup comparison analysis was performed to determine if both groups were comparable. Pearson’s chi-square or Fisher exact for categorical variables, while Student’s t-test or Mann-Whitney U test for independent continues variables. Values were presented as absolute and relative (percentage) or as median with interquartile range. A Mantel log-rank test to time to recovery until day-28 was conducted to compare the median time to recovery between groups and were stratified by the worst ordinal Scale during patient’s stay in the hospital[13,23]. Cox proportional hazard ratio was conducted to determine the recovery rate ratio (hazard ratio for recovery). Logistic regression was performed to determine the likelihood for mortality. P values < 0.05 were considered statistically significant. Statistical analysis was conducted using IBM SPSS Statistics 27 (IBM Corp., Armonk, NY, USA) software.

### Results

Of the 584 patients hospitalized with COVID-19 pneumonia from July 1, 2020, to Feb 20, 2021, 175/584 (29.9%) patients requiring 5 or more liters of supplemental O2 were screened for the study. 36/175 (20.5%) had some exclusion criteria. 51/139 (36.7%) patients admitted July 1 to December 31, 2020, received remdesivir with dexamethasone (standard therapy) and fulfilled eligibility for being “controls”. 88/139 (63.3%) eligible patients admitted from January 1 to February 20, 2021 received baricitinib with remdesivir and dexamethasone and were considered as “cases” (Fig 1).

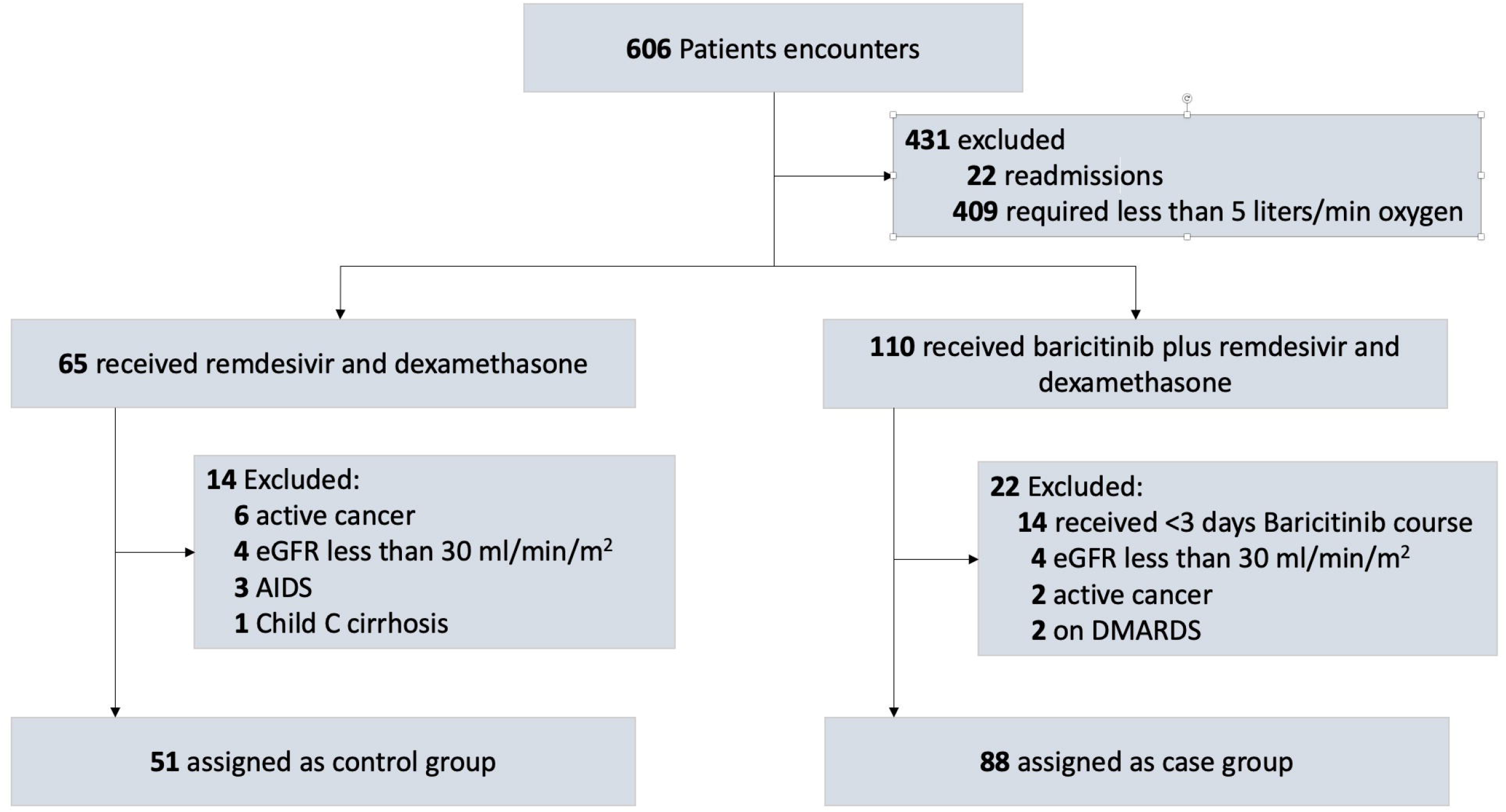

### Baseline Characteristics, severity, and clinical course

Both groups were comparable with respect to baseline characteristics, including demographics, comorbidities, and disease severity on admission, as shown in “Table 1”. The median age was 63 years with 56.1% being male and predominant Latinx (74.1%) followed by African-Americans (18.0%). Among the comorbidities, hypertension was common (56.8%), followed by Diabetes Mellitus (48.9%), Asthma (17.3%), and COPD (9.4%). The overall median Charlson Comorbidity Index was 3 88.4% of the cohort had severe COVID-19 (Ordinal scale 5 and 6) on admission, with 87.5% in the baricitinib group and 90% in the controls. Among the 10 patients requiring mechanical ventilation on admission (ordinal scale 7), 8 were in the baricitinib group, while only 2 were in the controls. The median time from symptoms onset to hospital admission was 6 days in both subsets. All patients in the cohort had moderate ARDS on admission. Calculated SOFA score within 24 hours of endotracheal intubation or institution of HFNC or the highest oxygen demand during the hospital course was 2 (IQR, 2-3), with similar distribution in both groups.

**Table 1.**
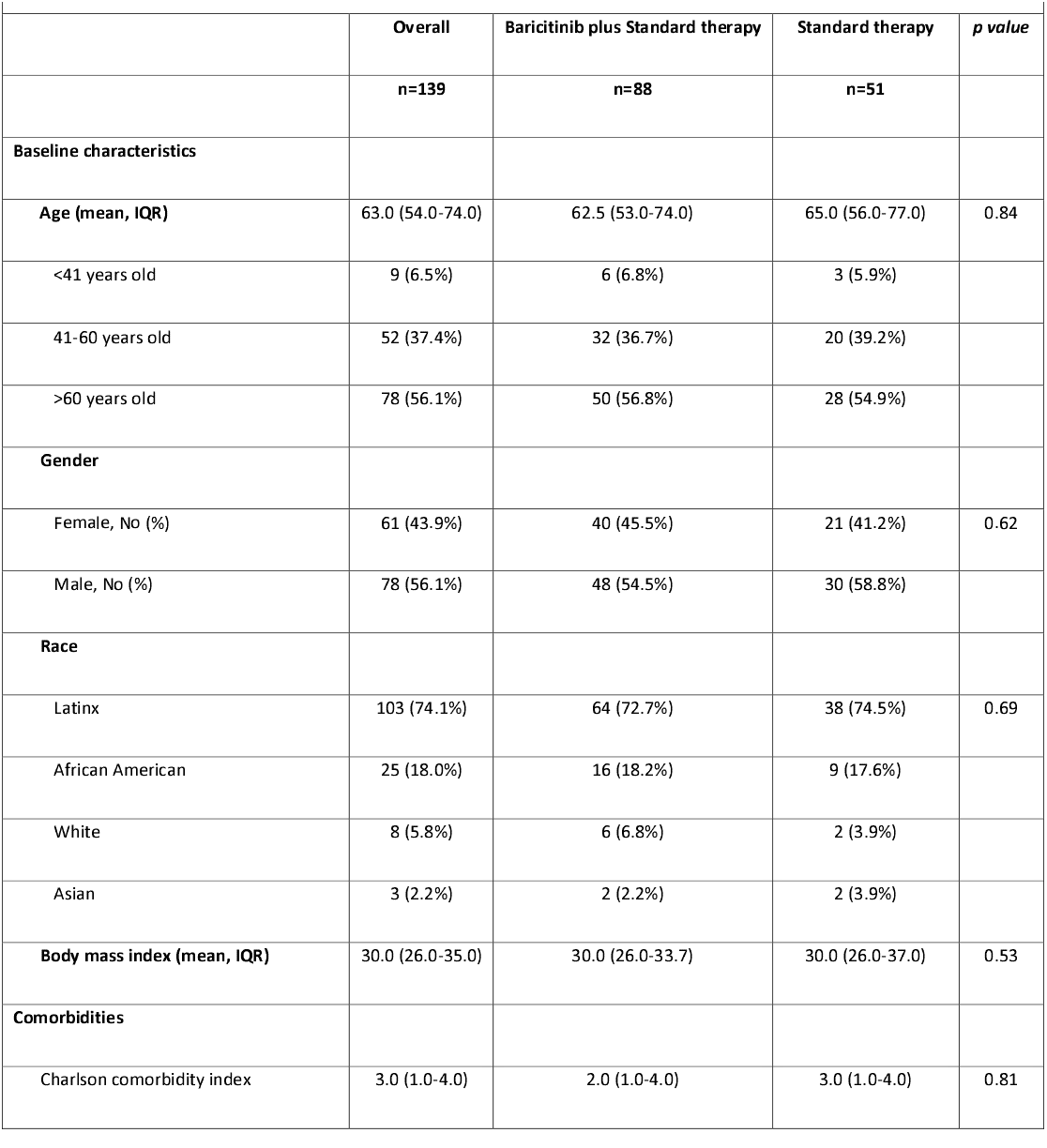
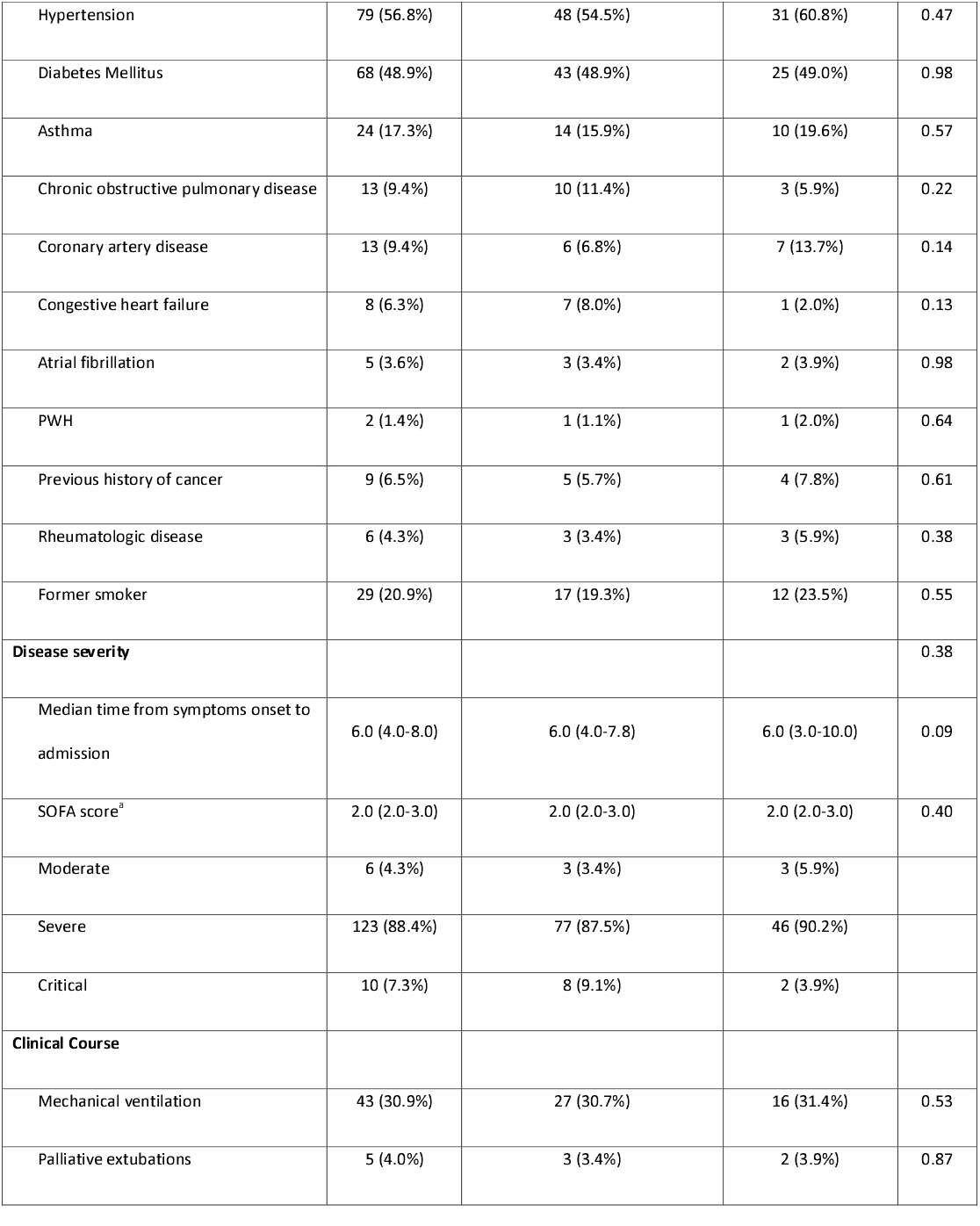
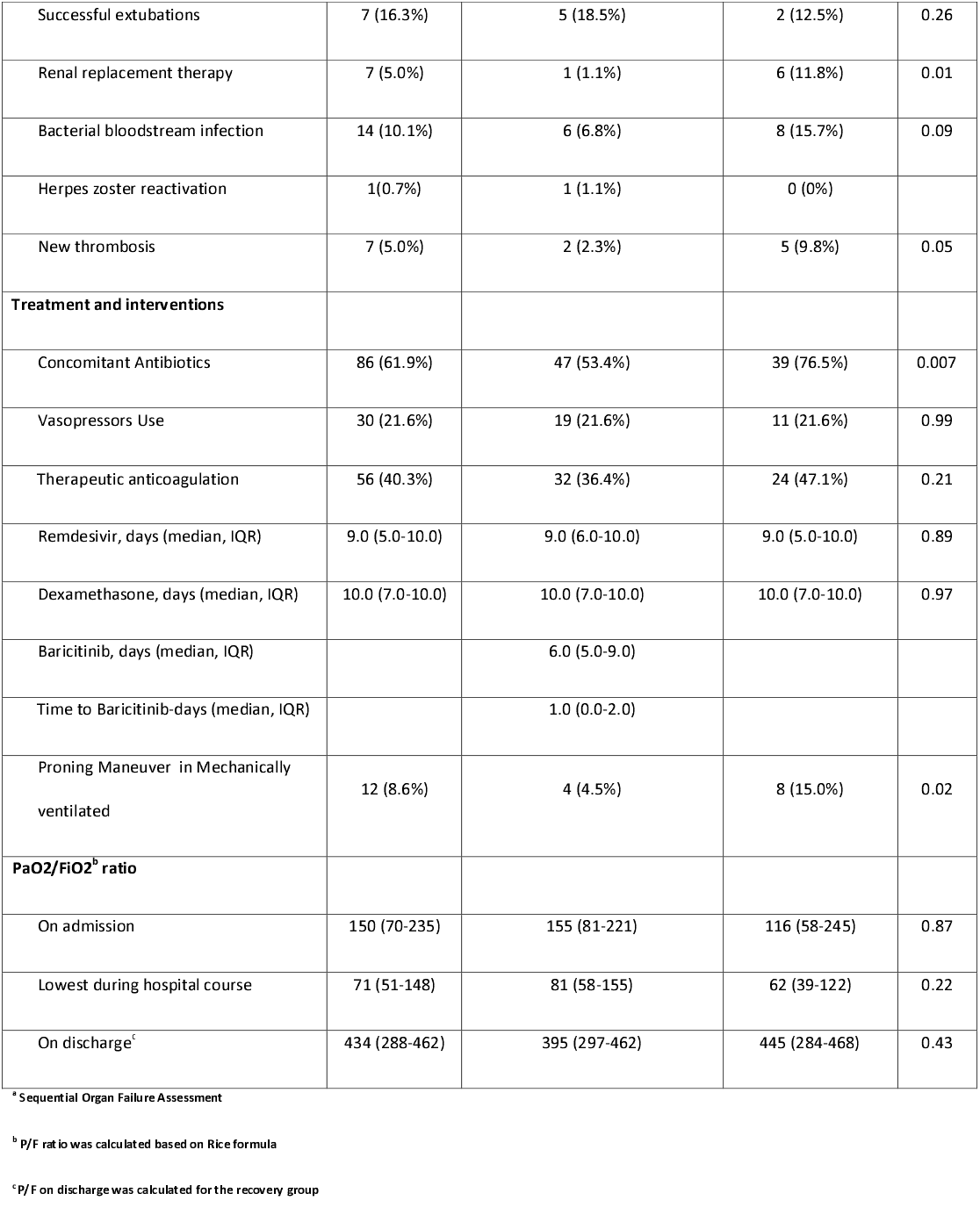
Baseline characteristics, clinical course, and treatment of Covid-19 patients treated either with Standard therapy or Baricitinib plus standard therapy.

|  |  |  |  |  |
| --- | --- | --- | --- | --- |
|  | **Overall** | **Baricitinib plus Standard therapy** | **Standard therapy** | ***p value*** |
| **Baseline characteristics** | **n=139** | **n=88** | **n=51** |  |
| **Age (mean, IQR)** | 63.0 (54.0-74.0) | 62.5 (53.0-74.0) | 65.0 (56.0-77.0) | 0.84 |
| <41 years old | 9 (6.5%) | 6 (6.8%) | 3 (5.9%) |  |
| 41-60 years old | 52 (37.4%) | 32 (36.7%) | 20 (39.2%) |  |
| >60 years old | 78 (56.1%) | 50 (56.8%) | 28 (54.9%) |  |
| **Gender** |  |  |  |  |
| Female, No (%) | 61 (43.9%) | 40 (45.5%) | 21 (41.2%) | 0.62 |
| Male, No (%) | 78 (56.1%) | 48 (54.5%) | 30 (58.8%) |  |
| **Race** |  |  |  |  |
| Latinx | 103 (74.1%) | 64 (72.7%) | 38 (74.5%) | 0.69 |
| African American | 25 (18.0%) | 16 (18.2%) | 9 (17.6%) |  |
| White | 8 (5.8%) | 6 (6.8%) | 2 (3.9%) |  |
| Asian | 3 (2.2%) | 2 (2.2%) | 2 (3.9%) |  |
| **Body mass index (mean, IQR)** | 30.0 (26.0-35.0) | 30.0 (26.0-33.7) | 30.0 (26.0-37.0) | 0.53 |
| **Comorbidities** |  |  |  |  |
| Charlson comorbidity index | 3.0 (1.0-4.0) | 2.0 (1.0-4.0) | 3.0 (1.0-4.0) | 0.81 |
| Hypertension | 79 (56.8%) | 48 (54.5%) | 31 (60.8%) | 0.47 |
| Diabetes Mellitus | 68 (48.9%) | 43 (48.9%) | 25 (49.0%) | 0.98 |
| Asthma | 24 (17.3%) | 14 (15.9%) | 10 (19.6%) | 0.57 |
| Chronic obstructive pulmonary disease | 13 (9.4%) | 10 (11.4%) | 3 (5.9%) | 0.22 |
| Coronary artery disease | 13 (9.4%) | 6 (6.8%) | 7 (13.7%) | 0.14 |
| Congestive heart failure | 8 (6.3%) | 7 (8.0%) | 1 (2.0%) | 0.13 |
| Atrial fibrillation | 5 (3.6%) | 3 (3.4%) | 2 (3.9%) | 0.98 |
| PWH | 2 (1.4%) | 1 (1.1%) | 1 (2.0%) | 0.64 |
| Previous history of cancer | 9 (6.5%) | 5 (5.7%) | 4 (7.8%) | 0.61 |
| Rheumatologic disease | 6 (4.3%) | 3 (3.4%) | 3 (5.9%) | 0.38 |
| Former smoker | 29 (20.9%) | 17 (19.3%) | 12 (23.5%) | 0.55 |
| **Disease severity** |  |  |  | 0.38 |
| Median time from symptoms onset to admission | 6.0 (4.0-8.0) | 6.0 (4.0-7.8) | 6.0 (3.0-10.0) | 0.09 |
| SOFA score<sup>a</sup> | 2.0 (2.0-3.0) | 2.0 (2.0-3.0) | 2.0 (2.0-3.0) | 0.40 |
| Moderate | 6 (4.3%) | 3 (3.4%) | 3 (5.9%) |  |
| Severe | 123 (88.4%) | 77 (87.5%) | 46 (90.2%) |  |
| Critical | 10 (7.3%) | 8 (9.1%) | 2 (3.9%) |  |
| **Clinical Course** |  |  |  |  |
| Mechanical ventilation | 43 (30.9%) | 27 (30.7%) | 16 (31.4%) | 0.53 |
| Palliative extubations | 5 (4.0%) | 3 (3.4%) | 2 (3.9%) | 0.87 |
| Successful extubations | 7 (16.3%) | 5 (18.5%) | 2 (12.5%) | 0.26 |
| Renal replacement therapy | 7 (5.0%) | 1 (1.1%) | 6 (11.8%) | 0.01 |
| Bacterial bloodstream infection | 14 (10.1%) | 6 (6.8%) | 8 (15.7%) | 0.09 |
| Herpes zoster reactivation | 1(0.7%) | 1 (1.1%) | 0 (0%) |  |
| New thrombosis | 7 (5.0%) | 2 (2.3%) | 5 (9.8%) | 0.05 |
| **Treatment and interventions** |  |  |  |  |
| Concomitant Antibiotics | 86 (61.9%) | 47 (53.4%) | 39 (76.5%) | 0.007 |
| Vasopressors Use | 30 (21.6%) | 19 (21.6%) | 11 (21.6%) | 0.99 |
| Therapeutic anticoagulation | 56 (40.3%) | 32 (36.4%) | 24 (47.1%) | 0.21 |
| Remdesivir, days (median, IQR) | 9.0 (5.0-10.0) | 9.0 (6.0-10.0) | 9.0 (5.0-10.0) | 0.89 |
| Dexamethasone, days (median, IQR) | 10.0 (7.0-10.0) | 10.0 (7.0-10.0) | 10.0 (7.0-10.0) | 0.97 |
| Baricitinib, days (median, IQR) |  | 6.0 (5.0-9.0) |  |  |
| Time to Baricitinib-days (median, IQR) |  | 1.0 (0.0-2.0) |  |  |
| Prone Maneuver in Mechanically ventilated | 12 (8.6%) | 4 (4.5%) | 8 (15.0%) | 0.02 |
| **PaO<sub>2</sub>/FiO<sub>2</sub><sup>b</sup> ratio** |  |  |  |  |
| On admission | 150 (70-235) | 155 (81-221) | 116 (58-245) | 0.87 |
| Lowest during hospital course | 71 (51-148) | 81 (58-155) | 62 (39-122) | 0.22 |
| On discharge<sup>c</sup> | 434 (288-462) | 395 (297-462) | 445 (284-468) | 0.43 |

<sup>a</sup> Sequential Organ Failure Assessment
<sup>b</sup> P/F ratio was calculated based on Rice formula
<sup>c</sup> P/F on discharge was calculated for the recovery group

### Clinical Course

Of the 33 patients who newly required mechanical ventilation during the course of hospitalization, 14 (87.5%) received standard therapy, while 19 (70.3%) received baricitinib with remdesivir and dexamethasone, as shown in “table 2”. The use of vasopressors was comparable between the two groups. Only 7 (16.3%) patients were successfully extubated, 5 (18.5%) in the case group and 2 (12.5%) among the controls. Six patients in the standard therapy cohort required new renal replacement therapy in comparison to one patient from the baricitinib group, with all of the seven patients requiring mechanical ventilation. The incidence of bloodstream infections in the total cohort was 10%, with 6.8% in the baricitnib group and 15.7% in the controls. Only two of the seven patients who had new thrombosis in the cohort received baricitinib. Use of concomitant antibiotics was higher in the standard therapy group in comparison with the baricitinib group (53.4%. vs. 76.5%, p=0.007). Among the 56 (40.3%) patients who received therapeutic anticoagulation, 32 (36.4%) were in the baricitnib group, while 24 (47.1%) were among the controls. The median number of days patients received dexamethasone was 10 days and remdesivir was 9 days. Patients in the baricitinib treatment group received the drug for a median duration of 6 (IQR, 5-9) days with a median time from admission of 1 (IQR, 0-2) day.

**Table 2.** Ordinal scale progression, time to recovery and mortality of Covid-19 patients treated either with Standard therapy or Baricitinib plus standard therapy.

|  | **Overall**<br>**n=139** | **Baricitinib plus Standard therapy**<br>**n=88** | **Standard therapy**<br>**n=51** | ***p value*** |
| --- | --- | --- | --- | --- |
| **Ordinal Scale on admission** |  |  |  | 0.38 |
| 4 (no oxygen requirement) | 6 (4.3%) | 3 (3.4%) | 3 (5.9%) |  |
| 5 (low flow oxygen device) | 75 (54.0%) | 50 (56.8%) | 25 (46.05) |  |
| 6 (high flow nasal cannula or NIV) | 48 (34.5%) | 27 (30.7%) | 21 (41.2%) |  |
| 7 (Mechanical ventilation) | 10 (7.2%) | 8 (9.1%) | 2 (3.9%) |  |
| **Higher Ordinal scale during hospital stay** |  |  |  | 0.79 |
| 5 (low flow oxygen device) | 20 (14.4%) | 14 (15.9%) | 6 (11.8%) |  |
| 6 (high flow nasal cannula or NIV) | 76 (54.7%) | 47 (53.4%) | 29 (56.9%) |  |
| 7 (Mechanical ventilation) | 43 (30.9%) | 27 (30.7%) | 16 (31.4%) |  |
| **Recovery status** |  |  |  |  |
| **Number of recoveries, overall** | 100 (71.9%) | 66 (75.0%) | 34 (66.7%) | 0.29 |
| Discharge without oxygen requirement | 58 (41.7%) | 34 (38.6%) | 24 (47.1%) |  |
| Newly prescribed home oxygen therapy on discharge | 42 (30.2%) | 32 (36.4%) | 10 (19.6%) |  |
| Median recovery time, days (95%CI)<sup>a</sup> | 12.0 (11.0-13.0) | 11.0 (10.0-12.0) | 14.0 (12.0-16.0) | 0.19 |
| Rate ratio (95% CI)<sup>b</sup> |  | 1.28(0.84-1.94) |  | 0.24 |
| **Number of recoveries, ordinal scale 5** | 20 (100%) | 14 (100%) | 6 (100%) |  |
| Median recovery time, days (95% CI)<sup>a</sup> | 9.0 (7.2-10.8) | 7 (4.8-9.2) | 11 (10.4-11.6) | 0.01 |
| Rate ratio (95% CI)<sup>b</sup> |  | 2.95 (0.053) |  | 0.04 |
| **Number of recoveries, ordinal scale 6** | 70 (95.9%) | 47 (100%) | 26 (89.7%) |  |
| Median recovery time, days (95% CI)<sup>a</sup> | 11.0 (10.2-11.8) | 10.0 (9.0-11.3) | 12.0 (11.3-12.7) | 0.01 |
| Rate ratio (95% CI)<sup>b</sup> |  | 1.80 (1.09-2.98) |  | 0.02 |
| **Number of recoveries, ordinal scale 7** | 7 (16.3%) | 5 (18.5%) | 2 (12.5%) |  |
| Median recovery time, days (95% CI)<sup>a</sup> | NE | NE | NE |  |
| Rate ratio (95% CI)<sup>b</sup> |  | 1.09 (0.21-5.63) |  | 0.91 |
| **Mortality** |  |  |  |  |
| Deceased (n, %) | 30 (21.6%) | 14 (15.9%) | 16 (31.4%) | 0.03 |
| Ordinal scale 5 | 0 (0%) |  |  |  |
| Ordinal scale 6 | 3 (3.9%) | 0 (0%) | 3 (10.3%) | 0.05 |
| Ordinal scale 7 | 27 (62.8%) | 14 (51.9%) | 13 (81.3%) | 0.05 |
| Odds ratio (95% CI)<sup>c</sup> |  | 0.41 (0.18-0.94) |  | 0.03 |

NE: Not Estimable
<sup>a</sup> Kaplan Meier survival estimates was conducted
<sup>b</sup> Cox proportional Hazard ration was performed
<sup>c</sup> Logistic regression analysis

### Primary Outcome

Baricitinib with remdesivir and dexamethasone reduced the median time to recovery by 3 days compared to the remdesivir-dexamethasone group for the total cohort. The median time to recovery was 11 days in the baricitinib treated group and 14 days in the control group, with rate ratio for recovery of 1.28 (95%CI 0.84-1.94, p=0.19), as shown in “table 2”. In the subgroup analysis by ordinal scale, a significant reduction in time to recovery was observed in patients with OS 5 and 6 among patients treated with baricitinib. Patients in OS 5 had a significant reduction in the median time to recovery by 4 days if they were treated with baricitinib (7 days) in comparison with control group (11days), rate ratio for recovery of 2.95 (95%CI 1.03-8.42, p=0.04). The median time to recovery among patients with OS 6 was significantly lower at 10 days in the baricitnib group compared to 12 days in the control group, with rate ratio for recovery of 1.80 (95%CI 1.09-2.98, *p*=0.02). The median time to recovery for patients requiring mechanical ventilation could not be calculated due to the small number of patients who recovered in both groups (Fig 2).

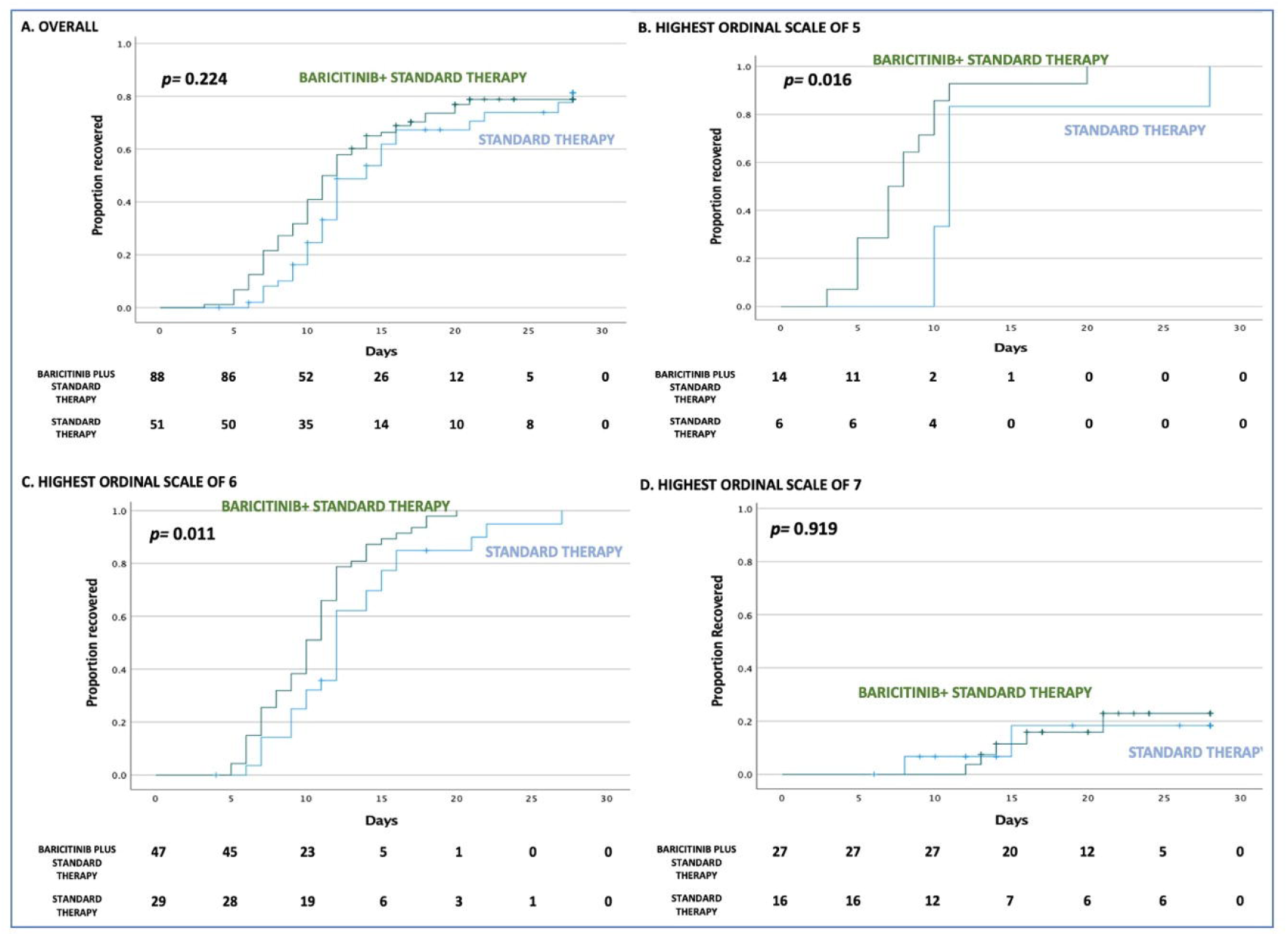
Cumulative recovery estimates are displayed in the overall cohort (panel A), in patients with ordinal scale of 5 (low flow oxygen device, panel B), in those with ordinal scale of 6 (High flow nasal cannula or NIV, panel C) and those with ordinal scale of 7 (mechanical ventilation, panel D).

### Secondary outcomes

Overall, 71.9% of patients in the study recovered; 75.0% in the baricitinib treated group and 66.7% in the control group. The changes in P/F ratio from hospitalization to discharge was greater in the baricitinib group in ordinal scale 5 and 6 (Fig 3). Of those who recovered in the baricitinib treated group, 36.4% required home oxygen therapy at discharge in comparison to 19.6% in the standard care group. The overall mortality rate was at 21.6%, with a significant reduction at 15.9% in the baricitinib treated group vs. 31.4% in the control group (p= 0.03). The likelihood of mortality was also noted to be significantly reduced in the baricitinib group (OR 0.41, 95%CI, 0.18-0.94, *p*=0.03). The survival rate by ordinal scale stratification, was 48.1% in OS 7 in the baricitnib group and 18.7% in the control care, with a difference of 29.4% (*p*=0.05), as shown in “table 2”.

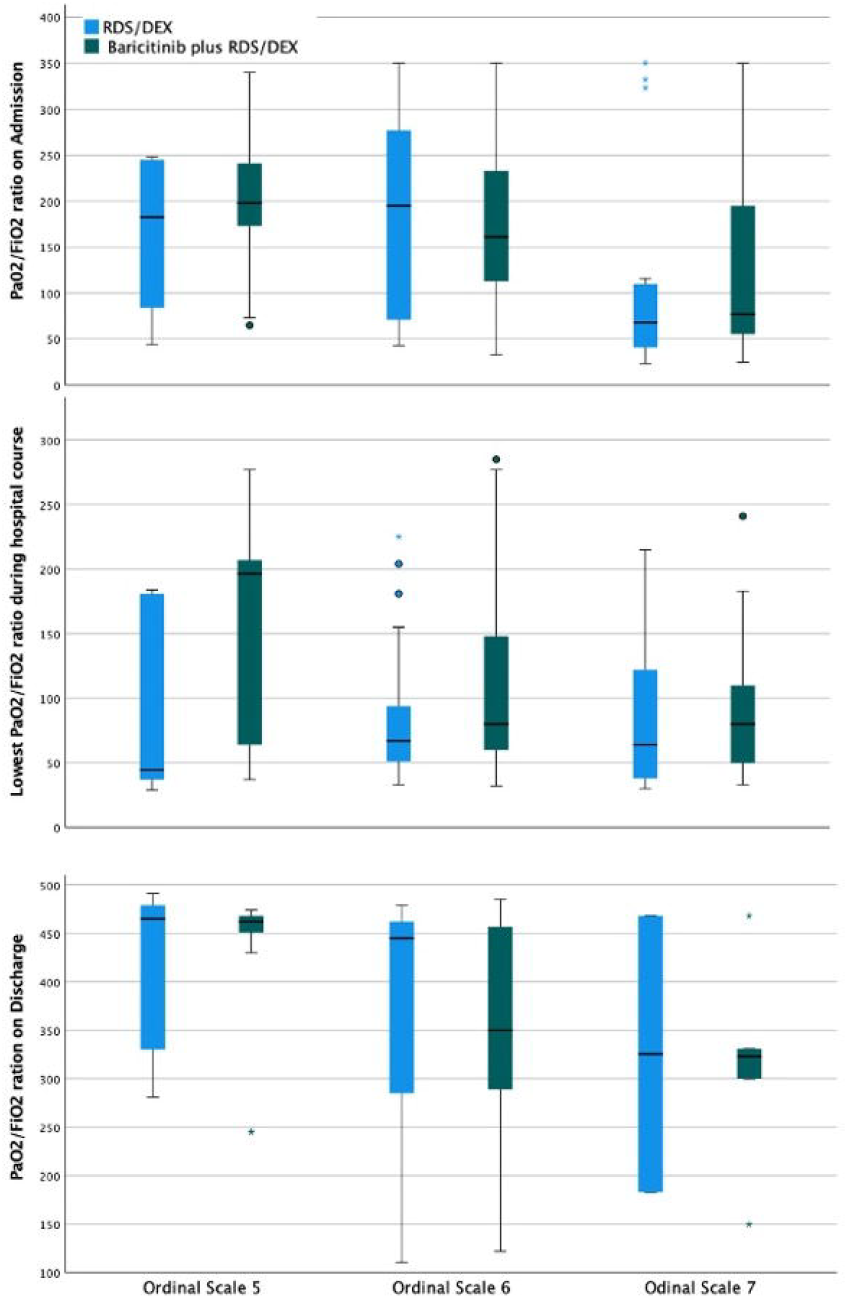
A: PaO2/FiO2 ratio on admission. B: Lowest Ordinal Scale during hospital course. C: Ordinal Scale on discharge among those who recovered

## Discussion

Corticosteroids play an important role as immunomodulator regulating the Cytokine release syndrome; The RECOVERY trial put in evidence that dexamethasone lower mortality rate in cases of severe and critical Covid-19 pneumonia and it became standard of care, others corticosteroids has been demonstrated survival benefit and improvement in respiratory parameters as well[9,11]. There is presently a lack of evidence on the impact of the combination of baricitinib with remdesivir and dexamethasone in the treatment of hospitalized patients with COVID-19 pneumonia[13]. In our retrospective observational study of patients with COVID-19 pneumonia admitted in OS 5 and 6, we observed a significant reduction in days to recovery when patients received baricitinib with remdesivir and dexamethasone in comparison to standard therapy. This benefit was not seen in the mechanically ventilated COVID-19 subgroup. The addition of baricitinib significantly reduced the rate and likelihood of mortality compared to the controls in the entire cohort. The 29% higher survival in the baricitinib group among the mechanically ventilated patients is noteworthy. An important highlight of the study and results are that the benefits were demonstrated in our predominantly minority population, including Latinx and African-American, who have been disproportionately affected by this pandemic. These results are particularly significant, as they expand our treatment options of severe and critical COVID-19 infections, with improved outcomes and recovery times translating to a reduced burden of the pandemic on healthcare. Dexamethasone became standard of care after The RECOVERY trial demonstrate mortality reduction for patient in the invasive mechanical ventilation and oxygen supplementation. The overall mortality rate in patient who received dexamethasone was 22.9%[9]. In the present study the use of baricitinib with remdesivir and dexamethasone therapy reduces mortality rate when is compared with standard of care therapy. We found the overall mortality rate to be 15.9% in the triple therapy group.

The research in therapeutics for COVID-19 has acknowledged that mitigating the immune response and preventing a hyperinflammatory state will improve clinical outcomes in patients with severe and critical infection. A randomized, double-blind, placebo-controlled trial (ACTT-2) showed that combination treatment with the JAK2 inhibitor baricitinib and remdesivir was superior to remdesivir alone in improving the time to recovery and clinical status in patients hospitalized with COVID-19 pneumonia[13]. An improvement in respiratory function was demonstrated following a short course of treatment with baricitinib and corticosteroid in patients hospitalized with moderate to severe ARDS due to COVID-19 pneumonia[16]. In a multicenter study of patients with moderate COVID-19 infection, Cantini et al. showed a mortality benefit, reduced ICU admission, and improved pulmonary function parameters in the group treated with Baricitinib plus lopinavir/ritonavir[24]. While there have been multiple small studies evaluating the impact of baricitinib with lopinavir/ritonavir, hydroxychloroquine, steroids in various combinations and doses[24,25], to the best of our knowledge, our study is the first to demonstrate a beneficial effect of combination of baricitnib with remdesivir and dexamethasone in reducing the time to recovery in patients hospitalized with severe COVID-19 infection, with overall mortality benefit in severe and critical infection.

The logic supporting the use of baricitinib includes diminishing the host inflammatory response and limiting the exuberate cytotoxic tissue damage. Baricitinib has two proven mechanisms of action. Firstly, it is a selective inhibitor of JAK1/JAK2, downregulating the STAT pathway and preventing the transducing signals for multiple cytokines[25–27]. The worsening trend of IL-6 cytokine concentration correlates with ARDS, organ injury, and mortality, highlighting the importance of implementing therapeutic options targeting this pathway. Baricitinib has a strong effect in reducing IL-6 and other proinflammatory cytokines in patients with COVID-19 infection and other diseases[24, 25]. The antiviral effect sustained its second mechanism of action, decreasing the rate of endocytosis of the virus by inhibiting the AP2-associated kinase and cyclin G-associated kinase that mediates the cellular membrane’s coating process after the virus binds the spike protein of the angiotensin-converting enzyme[28, 29].

Notably, we observed a lower incidence of bloodstream infections or new thrombosis among patients receiving baricitinib compared to those on standard therapy. Serious adverse events related to the drug were notably absent, and there was no instance of discontinuation of baricitinib due to side effects in the case group. Long-term use of baricitinib is associated with increased thrombosis incidence and viral/bacterial superinfections in patients with resistant Rheumatoid arthritis[28]. The safety profile of Baricitinib was assessed in the ACTT-2 trial, and the incidence of thrombosis and bacterial infections for patients with COVID-19 infections were similar in the baricitinib and placebo-group. Though the dual effect on immunosuppression with the concomitant use of baricitinib and dexamethasone raises concern for its additive risk of infection, our study failed to support these concerns, likely due to the shorter duration of use of baricitinib and faster rates of recovery reducing the days of therapy.

Our study had several limitations; firstly, this retrospective observational study included two groups of hospitalized COVID-19 pneumonia patients from two different timeframes (July-Dec 2020 and Jan-Feb 20, 2021). Though these groups had a similar distribution in demographic, baseline characteristics, and burden of the disease, we accept that there were confounders including “variants of concern” and mortality differences due to comparatively lesser burden of disease in the July-Dec timeframe. The study is also limited by sample size and generalizability. There was no standard length of therapy for the baricitinib-treated group and ranged from 3 to 10 days. The decision to stop baricitinib was at the discretion of the treating physician based on the patient’s clinical improvement. Thirdly we had two patients still hospitalized on mechanical ventilation, without a clear outcome, when the data was analyzed. Fourthly, the inherent limitations of retrospective studies, especially lack randomization, cannot be ignored, though we believe our encouraging results contribute to the therapeutic options for hospitalized severe COVID-19 patients till further robust data/evidence is available. Lastly, the lack of benefit in patients requiring mechanical ventilation could be due to the smaller sample size.

## Conclusion

Baricitinib with remdesivir and dexamethasone reduced time to recovery in patients with severe COVID-19 pneumonia requiring low flow oxygen and high flow oxygen or non-invasive ventilation, in comparison with standard care, including dexamethasone and remdesivir. Notably, this intervention did not demonstrate any benefit in time to recovery in critically ill patients requiring mechanical ventilation. Baricitinib use demonstrate mortality benefit in the overall analysis. The addition of baricitinib was not associated with an increase in bloodstream infections or new thrombosis.

## Data Availability

All data produced in the present study are available upon reasonable request to the authors

